# Interplay of Immunity, Climate, and Viral Evolution Explains Semiannual SARS-CoV-2 Dynamics with Implications for Control

**DOI:** 10.64898/2026.02.27.26347213

**Authors:** Samantha J. Bents, Kate M. Bubar, Hailey J. Park, Sophia T. Tan, Rachel E. Baker, Erin A. Mordecai, Nathan C. Lo

## Abstract

In the three years since Omicron emergence, SARS-CoV-2 dynamics have exhibited persistent twice-yearly waves in the United States, peaking in late summer and winter, with heterogeneity in timing and intensity across states. This semiannual pattern sharply contrasts with typical annual respiratory pathogen dynamics in the US, yet their underlying mechanisms and whether this pattern will persist remain poorly understood. Here, we tested several hypothesized mechanisms and found that a combination of waning immunity, climatic factors of relative humidity and temperature, variant activity, and vaccination captured divergent patterns in COVID-19 hospitalization incidence across 10 US states, from January 2022-November 2024. Applying a compartmental disease model, we identified that waning infection-derived immunity was the dominant driver of semiannual SARS-CoV-2 dynamics, with climate factors shaping the timing and magnitude of seasonal waves across US states. Scenario analyses indicated that if infection-derived immunity remains short in duration, semiannual dynamics influenced by climate are likely to persist, with attenuation in severe disease over time. In contrast, more durable infection-derived immunity, or a slower rate of immune-evading viral evolution, could lead to an epidemiologic transition to annual dynamics. In some states, summer waves approached the magnitude of winter waves, likely reflecting local climatic influences on transmission, suggesting that optimal vaccination strategies may vary by state. These findings have broad implications for understanding epidemic dynamics and informing vaccine policy, including seasonal timing and two-dose vaccine schedules for high-risk persons.

## Introduction

For three consecutive years following the emergence of Omicron in late 2021, SARS-CoV-2 transmission in the United States has been characterized by semiannual waves with substantial variation in timing and magnitude across US states^1^. These waves have consistently peaked in late summer and winter, suggesting that the observed patterns could reflect emerging endemic behavior. This semiannual dynamic contrasts with the majority of other endemic respiratory viruses, such as influenza and respiratory syncytial virus, which typically exhibit a single annual winter peak in temperate locations^1–3^. A key question for public health is whether these semiannual dynamics will be sustained over time or settle into an annual cycling pattern. Understanding the mechanisms driving these epidemic patterns, and how they produce state-level differences in timing and intensity of SARS-CoV-2 waves, is important for anticipating variation in disease burden and designing the optimal timing and vaccine schedule for high-risk individuals.

The seasonality of endemic pathogens is driven by the interplay between population immunity, pathogen evolution, climate conditions, human behavior and mobility, and control measures such as vaccination, which together shape characteristic patterns of disease circulation^2–6^. In the early phase of the COVID-19 pandemic (2020-2021), SARS-CoV-2 dynamics were dominated by population susceptibility and the emergence and spread of novel variants, with new variants often being associated with changes in infectiousness and/or pathogenicity^7,8^. Widespread vaccine rollout beginning in early 2021 further shaped population immunity, with uptake in subsequent years typically peaking in autumn across US states^9^. Over time, reinfections have become the predominant pattern in both vaccinated and unvaccinated populations, consistent with partial and waning protection from infection-acquired and vaccine-derived immunity, but with population-wide reduced clinical severity of disease^10,11^.

Beyond immunity, climatic drivers, such as humidity and temperature, modulate the seasonal transmission dynamics of other respiratory endemic pathogens such as influenza and respiratory syncytial virus^2,12,13^, but their role in shaping SARS-CoV-2 dynamics remains uncertain in the current epidemiologic era. Although laboratory studies have demonstrated that temperature and relative humidity affect SARS-CoV-2 virus viability in indoor environments^14^, population-level studies from the early pandemic suggested that associations between disease burden and climate were limited due to high population susceptibility amid rapid variant turnover^15,16^. Human behavior further interacts with these processes: early reductions in mobility and the introduction of nonpharmaceutical interventions reduced transmission^17^, but the return of typical mobility and travel patterns likely reintroduced additional behavioral impacts on transmission in subsequent years^18^. Evidence from other respiratory pathogens indicates that recurring behavioral patterns, such as school calendars and seasonal travel, can shape seasonal dynamics outside pandemic contexts^19–21^, suggesting that behavior may continue to contribute to the evolving dynamical cycling of SARS-CoV-2.

Here, we develop competing mathematical modeling frameworks to test hypotheses about which mechanisms capture the semiannual SARS-CoV-2 transmission dynamics and their variation across US states from 2022-2024. We evaluate the contribution of waning infection-acquired and vaccine-derived immunity, climatic drivers of temperature and relative humidity, variant activity, and behavioral factors. Our analysis has implications for US public health responses and vaccine policy, including whether to continue recommending a two-dose schedule for high-risk individuals to align with semiannual dynamics and how to optimize seasonal timing to reduce US COVID-19 burden.

## Results

### Model Comparison to Explain post-Omicron SARS-CoV-2 Dynamics in United States

Using a Bayesian calibration process, we fit seven competing mechanistic compartmental models with a Susceptible-Exposed-Infected-Recovered-Susceptible (SEIRS) structure with vaccination to observed weekly COVID-19 hospitalization incidence surveillance data from January 2022 to November 2024 across ten US states available in the COVID-NET surveillance system (see Methods and Supplementary Materials). Each model structure tested different hypotheses about the drivers of recent SARS-CoV-2 dynamics, and assessed whether inclusion of candidate mechanistic drivers of transmission improved the model agreement with observed disease patterns (**Table 1**). We found that incorporating rapid waning of infection- and vaccine-derived immunity, temperature- and relative humidity-dependent transmission, and modest increases in immune waning during periods of novel variant emergence (immune evasion) into a mechanistic model best explained SARS-CoV-2 dynamics (Model 6 in **Table 1**; **Figure 1A**). This model structure captured heterogeneous seasonal trends and magnitudes in COVID-19 hospitalization across the ten US states (**Figure 1B**).

**Figure 1.**
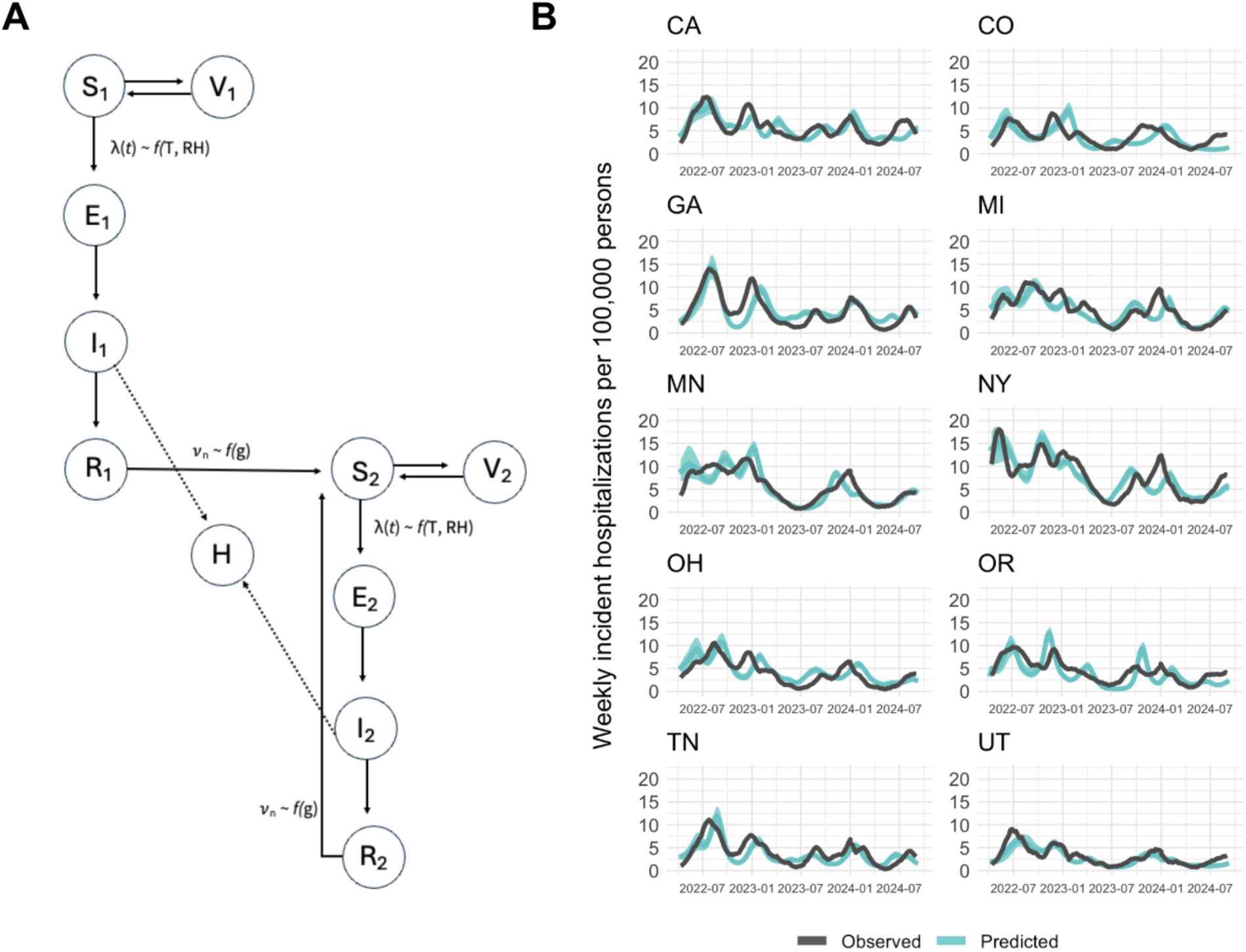
Mechanistic model structure incorporating waning immunity, climate-dependent transmission, and novel variant immune evasion captures SARS-CoV-2 dynamics across US states from January 2022-November 2024. **A)** Compartmental model structure of Susceptible-Exposed-Infected-Recovered-Susceptible (SEIRS) with vaccination (V) was used to simulate SARS-CoV-2 dynamics in the US. In this model, the force of infection *λ*(*t*) was a function of temperature (T) and relative humidity (RH). Upon first infection, individuals could be hospitalized (H). After the first infection, individuals recovered and developed immunity before eventually waning into a second set of compartments with the same structure, but with a reduced rate of hospitalization upon subsequent infection. During periods of novel variant emergence and spread (defined in Methods), the rate of waning *v_n_* increased by *f*(*g*) to account for immune evasion. The model schematic includes notations for how each transmission mechanism (see Table 1) was incorporated into the final selected model. Alternative model structures were evaluated (see Supplementary Materials). The model was initialized to January 2022 conditions, following Omicron emergence. **B)** Comparison of model estimated weekly COVID-19 hospitalization incidence (blue line, with 95% credible interval) using a model structure incorporating waning of infection-acquired and vaccine-derived immunity, climate factor-modulated transmission, and variant activity against observed weekly COVID-19 hospitalization incidence (black) in 10 US states based on the COVID-NET surveillance study.

**Table 1:** Comparison of model structures accounting for different transmission relevant mechanisms to explain SARS-CoV-2 dynamics in the United States from 2022-2024.

|  | Model 1 | Model 2 | Model 3 | Model 4 | Model 5 | Model 6 | Model 7 |
| --- | --- | --- | --- | --- | --- | --- | --- |
| **Assumption** |  |  |  |  |  |  |  |
| Infection- and vaccine-derived immunity and waning | ✓ | ✓ | ✓ | ✓ | ✓ | ✓ | ✓ |
| Temperature-dependent transmission |  | ✓ |  | ✓ | ✓ | ✓ | ✓ |
| Relative humidity-dependent transmission |  |  | ✓ | ✓ |  | ✓ | ✓ |
| Behavior-dependent transmission |  |  |  |  | ✓ |  | ✓ |
| Variants influence waning rate |  |  |  |  |  | ✓ | ✓ |
| **Model performance** |  |  |  |  |  |  |  |
| Log likelihood<sup>a</sup> | -16,880 | -16,718 | -14,179 | -13,989 | -16,347 | -13,060 | -13,331 |
| AIC | 33,772 | 33,450 | 28,374 | 27,996 | 32,708 | 26,140 | 26,684 |
| BIC | 33,805 | 33,488 | 28,418 | 28,045 | 32,746 | 26,195 | 26,744 |

<sup>a</sup>Final log likelihood values were adjusted for the relative population sizes across ten states in the calibration.

The estimated short-term infection-acquired immunity against infection (mean duration of 5.0 months (95% Credible interval (CrI): 4.7 5.2)) and vaccine-acquired immunity (mean duration of 3.1 months (CrI: 2.2, 5.2)), enabled semiannual dynamics, while climate covariates shaped the timing and magnitude of waves across states. During periods of high variant activity (defined as periods when a single variant exceeded 70% of sequences reported for >1 month), we estimated that the waning rate from natural infection increased by 24.8% (CrI: 14.2, 35.4), suggesting immune evasion associated with novel variant emergence (see Methods). A model validation analysis was conducted from December 2024 to January 2026, and demonstrated comparable trends between the model-predicted and observed severe COVID-19 cases over time (**Supplementary Figure S3).**

### Exploring Climatic Drivers of SARS-CoV-2 Transmission

Using our best fit model, we assessed the time-varying relative importance of mechanistic drivers of transmission. We regressed the model-fitted weekly effective reproduction number, R_e_(*t*), against key mechanistic drivers including population immunity (from infection-acquired or vaccine-derived immunity, also incorporating contribution of immune evasion due to novel variants), temperature, and relative humidity, for the 2022-23 and 2023-24 seasons. We found that changes in population immunity from both infection and vaccination combined explained the highest share of the variation in R_e_(*t*), accounting for an average of 85-89% of variation across seasons (**Fig 2A**), where the majority source of immunity was infection-derived (on average, the combined immune compartment was 98% individuals in recovered compared to vaccinated compartments). The influence of relative humidity variation on transmission peaked between the months of August-October, explaining a maximum of 9-15% of the variance in R_e_(*t*) across seasons. Temperature variation explained at most 12-25% of the variance in R_e_(*t*) between the months of November-February across states and seasons (**Fig 2B**).

**Figure 2.**
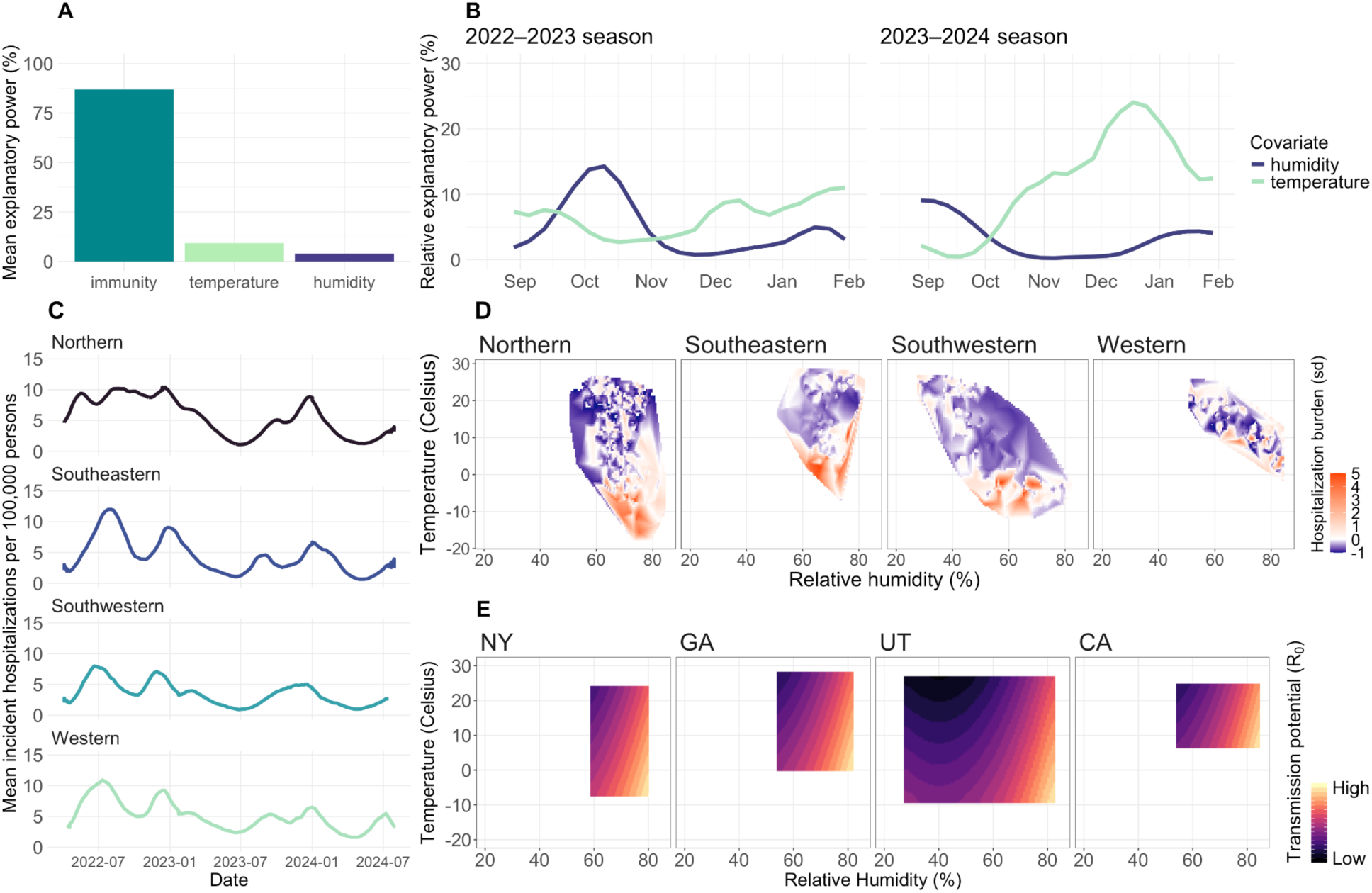
Estimating the contribution of mechanisms shaping SARS-CoV-2 transmission dynamics in the United States, including population immunity and climate factors of temperature and relative humidity. **A)** Mean relative importance of immunity (infection-acquired immunity, influenced by novel variant emergence, and vaccine-derived immunity), temperature, and relative humidity using the primary model fit. The relative importance was estimated by regressing model-fitted weekly R_e_(*t*) values against relative humidity, observed temperature, and fraction of the population that is immune, via both vaccine- and infection-derived immunity, in five-week intervals over the study period across all US states in the study. **C)** Time-varying relative importance of relative humidity and temperature across all states in the 2022-23 season (left) and the 2023-24 season (right), aggregated over all US states in the study, where the remainder of relative explanatory power sums to 100% and is attributed to immunity. **D)** Mean incident COVID-19 hospitalizations per 100,000 persons in Northern (MI, MN, NY, OH), Southwestern (CO, UT), Southeastern (GA, TN), and Western (CA, OR) US states from June 2022-November 2024. **D)** Heat maps displaying the relationship between observed temperature, observed relative humidity, and standardized hospitalization burden across four climatic regions. **E)** Model-estimated relationship between temperature, relative humidity, and transmission potential (R_0_) in four state case examples representing four unique climatic regions. The estimated climate-transmission relationship was fit globally across all states.

To assess how temperature and relative humidity may influence SARS-CoV-2 dynamics together, we aggregated US states into four broader geographic regions where divergent hospitalization time series patterns were observed: Northern states (MI, MN, NY, OH), Southeastern states (GA, TN), Southwestern states (CO, UT), and Western states (CA, OR) (**Figure 2C**). Empirical analyses demonstrated that higher COVID-19 hospitalization burden generally occurred at lower temperatures, while substantial disease burden was observed during periods of both high and low relative humidity (**Figure 2D**). To isolate the influence of climate factors on transmission, independent of underlying susceptible dynamics, we then estimated transmission potential (basic reproduction number, R_0_) across a range of observed temperature and relative humidity conditions. We used case examples from four US states, to probe how the globally fitted relationship between transmission and climate may influence disease dynamics in each of the broader geographic regions (**Figure 2E**). We found that the range of predicted R_0_ was highest in the Northern states, similar for the Western and Southeastern states, and lowest in the Southwestern states.

### Periodicity and Counterfactual Scenarios Perturbing Semiannual Dynamics

Consistent periodicity in recent transmission dynamics may suggest stable semiannual waves, whereas changes or variation in these intervals could indicate uncertainty in whether semiannual waves will persist. To assess the consistency of semiannual dynamics, we measured the timing between the peaks of the winter and summer wave of COVID-19 hospitalizations across states over time. We found that the timing of peak hospitalizations has remained generally consistent across states during the winter peaks (mean across states = week 50 and standard deviation (sd) = 3 for Winter 2022-23 and mean = week 52 with sd = 2 for Winter 2023-24), with the timing of summer waves being more variable between years but similarly consistent across states (mean = 31, 39, 34, sd = 4, 2, 3 for Summer 2022, 2023, and 2024, respectively) **(Figure 3A)**. However, differences in the number of weeks between the summer and winter peaks were observed across regions. Across seasons, the number of weeks between the summer and subsequent winter peak was the highest in the Southwestern region (mean = 21 weeks) and lowest in the Northern states (mean = 14 weeks), with the Western and Southeastern regions showing similar trends (mean = 19 and 17 weeks, respectively).

**Figure 3.**
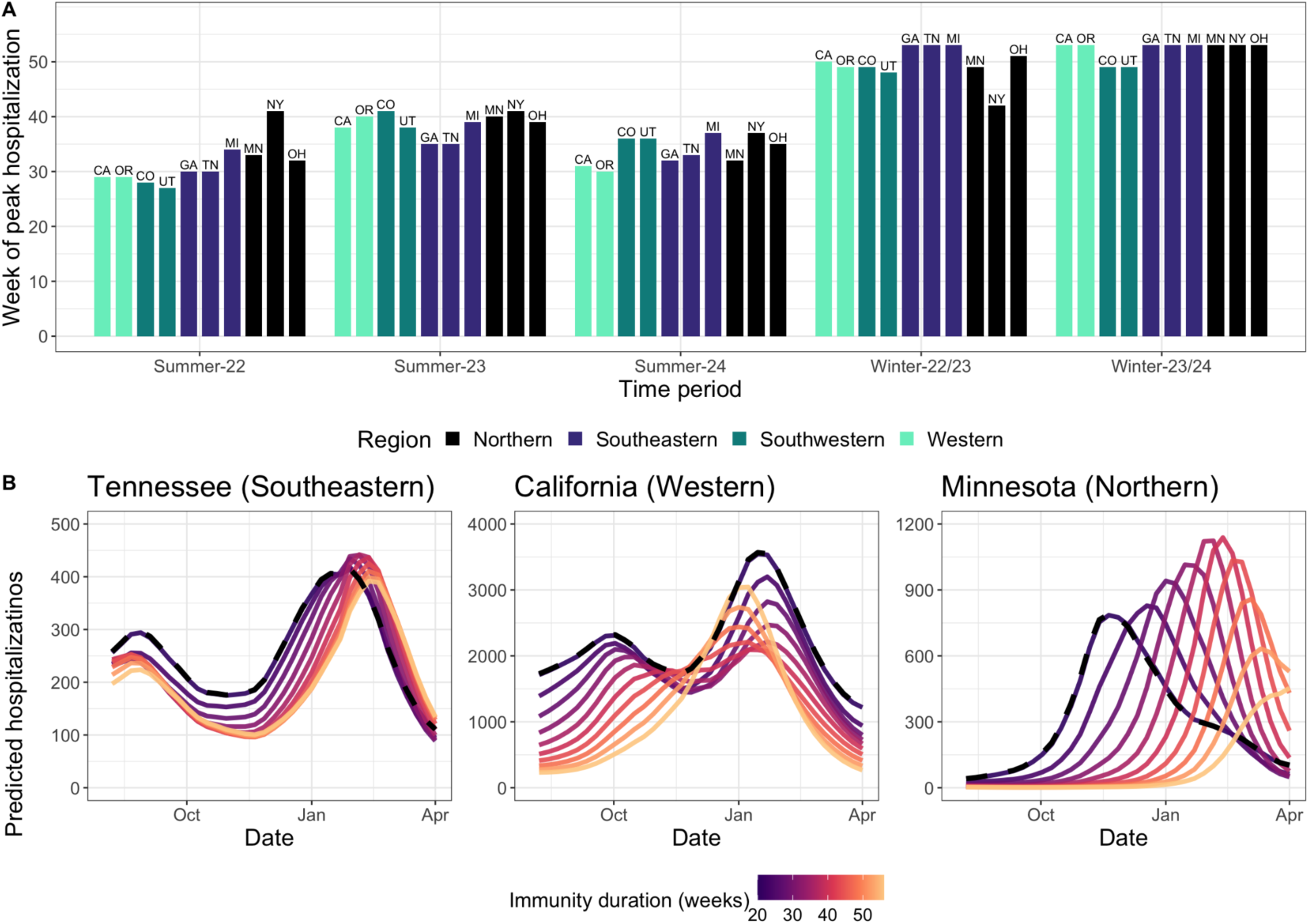
Scenario analysis of mechanisms capable of disrupting semiannual SARS-CoV-2 dynamics. **A)** Observed week of peak hospitalization by state and time period for Summer 2022-2024 and Winter 2022-2024, suggesting consistent timing of peaks across years, colored by region. **B)** Projected SARS-CoV-2 dynamics using the calibrated model (black dashed line) with scenarios testing varying longer durations of infection-acquired immunity (which could also represent a slower rate of immune evading viral evolution), ranging from 20 weeks (approximate calibrated estimate) to 52 weeks. We used three large US states with diverse SARS-CoV-2 dynamics as case examples: Tennessee (left), California (middle), and Minnesota (right). Scenarios with a longer infection-acquired duration of immunity were more likely to disrupt the semiannual dynamics.

To explore the mechanisms that could disrupt contemporary SARS-CoV-2 semiannual dynamics across broad US regions, we explored changing immunity and/or viral evolution scenarios. Using model fits, we implemented counterfactual scenarios from January 2023-June 2024 in three US states from different regions (Tennessee, California, and Minnesota) where longer durations of infection-acquired immunity were tested (which could also represent a slower rate of immune evading viral evolution). We found that increasing the duration of immunity supported a transition from semiannual dynamics to annual cycling patterns, particularly in California and Minnesota, with the peak of the predicted annual winter wave in Minnesota occurring several months later than that predicted for California (**Figure 3B**). In Tennessee, our model predicted more durable semiannual dynamics.

### Optimizing Vaccination Timing and Schedule by Accounting for States Differences in Seasonal Dynamics

Given the rapid waning of vaccine-derived protection against infection and state-by-state variation in timing of SARS-CoV-2 waves across the US, the optimal timing of vaccine uptake to minimize COVID-19 hospitalization burden may differ geographically. Over the 2023-2024 period, vaccine uptake consistently peaked in September-October across states, despite some differences in coverage, yet the relative magnitude of the summer and winter wave of COVID-19 hospitalizations differed substantially (**Figure 4A**). To assess the impact of vaccination timing by state, we simulated shifting observed vaccination timing between −6 and 6 weeks, and assessed the percent change in predicted hospitalizations in the 2023-24 season in these counterfactual scenarios (**Figure 4B**). We found that Western and Southeastern states showed increased benefit from relatively late vaccination uptake. Assessment of the relative cumulative size of hospitalizations in the summer versus winter outbreak revealed regional patterns, whereby in Northern States, the highest risk was concentrated during the winter, whereas in Southeastern and Western states, risk was more evenly distributed across the summer versus winter peak, motivating consideration of potential summer vaccination (**Figure 4C**). To compare seasonal targeting strategies across states, we estimated the relative impacts of annual fall (peaking in October), annual summer (peaking in August), and two-dose (summer and fall) vaccination strategies compared to a no vaccination scenario, using realistic (ranging from 9-27% cumulative coverage across states, in line with observed uptake) and optimistic vaccine coverage (**Figure 4D, Supplementary Figure S4**). In Western and Southeastern states, where summer and winter burdens were similar, fall versus summer annual vaccination had similar impact. In some states, such as Oregon, the summer dose provided greater reductions in hospitalizations than the fall dose. In other states with more concentrated winter peaks, fall vaccination had substantially greater impact than summer vaccination. A two-dose strategy was the most effective vaccine schedule across all states, but had a larger incremental impact in states with similar summer and winter peaks (**Figure 4D**).

**Figure 4.**
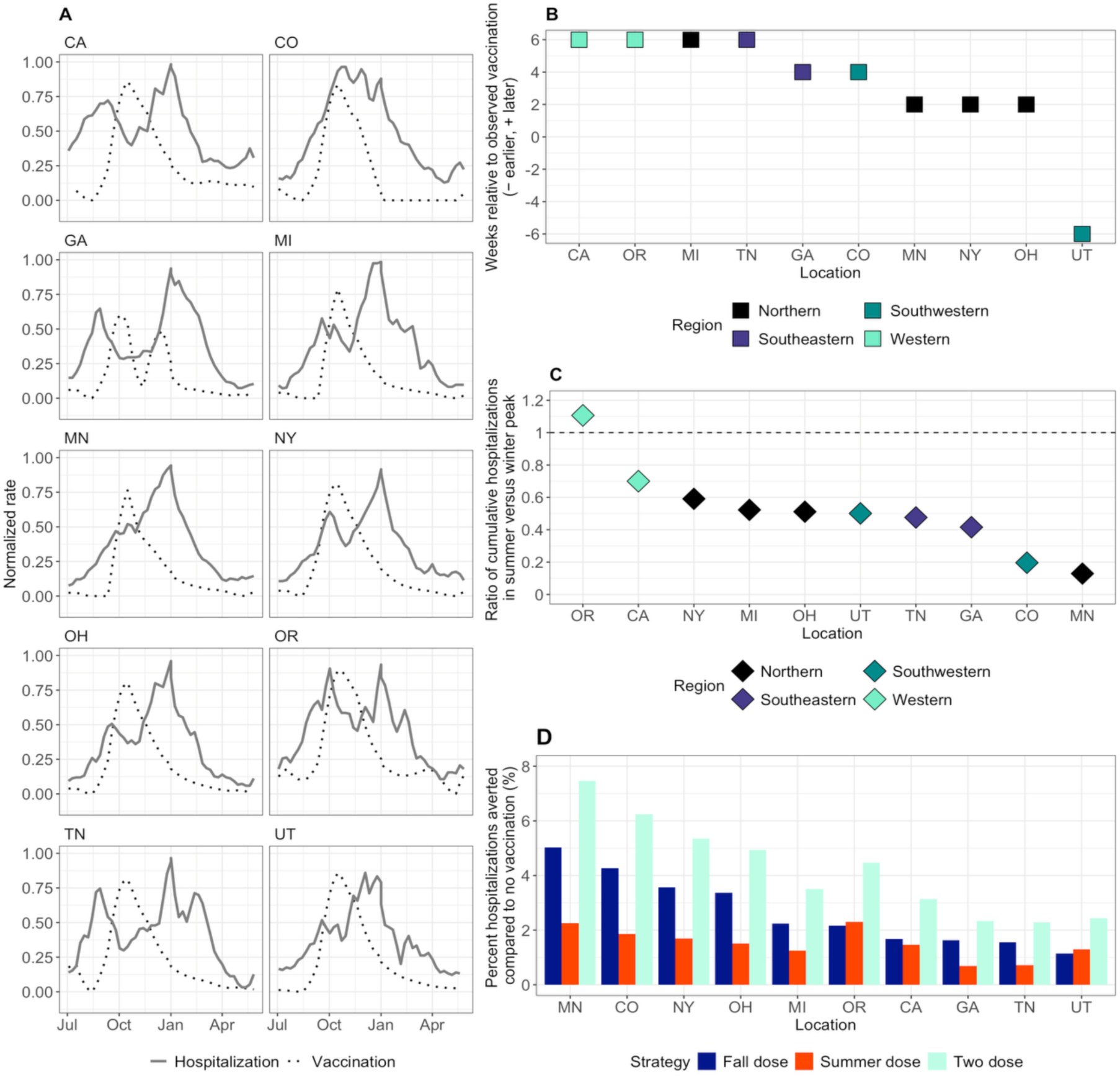
Implication of SARS-CoV-2 seasonal dynamics on optimal vaccination timing and schedule. **A)** Normalized observed vaccination rates (dotted lines) and COVID-19 hospitalization rates (solid lines) across US states June 2023-June 2024. **B)** Optimal shift in vaccination timing where vaccination curves were shifted earlier (negative direction) or later (positive direction) relative to observed timing (timing of vaccination peak = 0), colored by region. **C)** The relative magnitude of severe COVID-19 cases over winter versus summer peaks across states, suggesting regional clustering in the size of these peaks. Points above the dashed line (y =1) indicate a larger cumulative summer wave than winter wave. **D)** Percent change in predicted hospitalizations under an annual summer dose, annual fall dose, and two-dose vaccine schedule compared to no vaccination, which suggests there may be comparable impact for annual fall and summer dose administration in some states. We used observed annual state-specific coverage levels (ranging from 9-27% coverage), although tested more optimistic vaccine uptake in sensitivity analysis (Supplementary Figure S4). Since the model does not incorporate age and risk stratification, these estimates are an underestimate of the absolute impact of differing vaccination on COVID-19 hospitalization burden but the relative pattern will be conserved.

## Discussion

In this study, we identified that a mechanistic model accounting for waning immunity, climate-dependent transmission, and novel variants with immune evasion outperformed alternative models and captured the variability in semiannual SARS-CoV-2 dynamics across US states in the post-Omicron era. Our findings suggested that waning immunity from natural infection was the predominant driver of these atypical semiannual dynamics, with temperature and relative humidity shaping regional variation in timing and magnitude of peaks. Vaccine-derived immunity contributed less to dynamics due to lower uptake and modest vaccine efficacy and rapid waning of protection against infection. Scenario analyses suggested that if recent patterns of infection-acquired immunity duration continue, semiannual waves may persist and could imply a stable endemic state. In contrast, more durable immunity against infection, or a slower rate of immune-evading viral evolution, could support an epidemiologic transition to annual dynamics.

In our selected model, the predominant driver of semiannual SARS-CoV-2 dynamics was waning protection from infection-acquired immunity. However, it is difficult to disentangle waning immunity from immune evasion associated with viral evolution and novel variants; our parameterization of waning infection-acquired immunity likely represents a combination of these mechanisms. Our model estimates for the duration of protection following infection and vaccination aligned well with previously published estimates, providing further evidence for these mechanisms^22–25^. We also found that the inclusion of dominant variants leading to immune evasion (via increased rate of waning) improved the model fit, underscoring the role of novel variants. Periods of sustained variant dominance, coinciding with the BA.5, JN.1, and XBB1.5 variant emergence, likely fueled larger epidemic size when these variants were circulating by effectively placing more people at risk of infection who would have had protection against previous variants.

Our analysis suggests that temperature and relative humidity have an influential role in driving differential SARS-CoV-2 dynamics across US states, with a negative correlation with temperature driving winter peaks and a U-shaped relationship with relative humidity driving summer peaks. Importantly, these modeling findings are aligned with laboratory findings, which showed that the half-life of SARS-CoV-2 particles increased at low temperatures and at extreme low and high relative humidity^14^. At lower temperatures, the envelope of the virus increases in rigidity and viral proteins are more chemically stable, facilitating increased transmission potential^14,26^. Coronaviruses have been shown to display a nonlinear relationship with relative humidity, with virus particles more stable at both low and high levels^14,27^. The effects of relative humidity are postulated to be multifactorial; for example, at low relative humidity, SARS-CoV-2 particles aerosolize more quickly and remain suspended for longer periods of time, while human mucosal membranes are also typically drier, leading to increased virus infectivity^28,29^. At higher relative humidity, respiratory droplets remain viable longer due to being larger and settle more rapidly, supporting fomite-mediated transmission^14^. Our modeling analysis suggested that the relationship between SARS-CoV-2 transmissibility and these climate variables may be detectable at the population level.

While our findings support the role of climate in driving differential SARS-CoV-2 dynamics across the US, it is possible that these variables may be proxies for human behavioral patterns that follow seasonal trends. Prior studies have found that US populations are more likely to spend time indoors in winter months in Northern states, while in Southern states, indoor activity peaks in both winter and summer, likely a behavioral response to high outdoor humidity and heat^30^. Time spent indoors has been associated with increased probability of airborne transmission with SARS-CoV-2^31^, suggesting that climatic variables included in our modeling framework may also, to some extent, represent human behavior associated with increased time spent indoors. However, we tested the inclusion of a variable representing mobility that accounted for time spent indoors, which did not affect model performance or reduce the relation of climatic factors, supporting a more direct role of climate factors in transmission.

Although respiratory endemic pathogens such as influenza and RSV typically peak in the winter in the Northern Hemisphere, summer circulation is not entirely unique to SARS-CoV-2. Several studies have shown circulation of adenoviruses and under some circumstances, seasonal coronaviruses OC43 and HKU1, during the summer, particularly during unusually cool or humid summers, albeit typically at low levels relative to winter peaks^32–34^. These pathogens generally demonstrate slower immune-evading viral evolution compared to SARS-CoV-2, but may be driven by similar climatic conditions, particularly given high genetic similarities between beta coronaviruses OC43 and HKU1 and SARS-CoV-2^35^.

The future of SARS-CoV-2 dynamics, mainly whether semiannual patterns will persist, is a key question that our study aimed to address. While the overall magnitude of COVID-19 hospitalizations has declined over time, the semiannual patterns have remained consistent. Our findings suggest that continued short-term infection-derived immunity (and/or continued viral evolution with immune evasion) would sustain semiannual dynamics shaped by climate factors. However, if infection generates more durable protection, this could plausibly support an epidemiologic transition to an annual pattern or sustain low-level transmission patterns, where less pronounced seasonal patterns emerge. Counterfactual scenarios suggested that longer infection-derived immunity could have differential impacts across US regions, with Western and Northern states showing higher propensity for annual dynamics, while Southeastern states may persist longer with semiannual dynamics. In Northern states, we projected a later annual peak under a longer infection-acquired immunity duration scenario compared to Western states, suggesting that climate may play a role, with temperature being a potentially larger driver in Northern states while higher relative humidity may be more influential in Western states. Southeastern states observe relatively larger fluctuations in both temperature and relative humidity, which may improve durability of semiannual dynamics. The continued investment in SARS-CoV-2 surveillance data is necessary to monitor shifts in seasonal patterns.

Our model findings have implications for US vaccine policy. First, this study supports a two-dose schedule in high-risk patient populations to align with semiannual dynamics, especially in certain states with similar magnitudes of summer and winter COVID-19 waves^36,37^. Two doses of the current COVID-19 vaccine are currently recommended for individuals aged 65 years and older and individuals (age 6 months and older) who are moderately or severely immunocompromised, generally given 6 months after the first dose, although without specific guidance on timing^38^. Second, our findings also support some modest benefit of optimizing the timing of vaccination, currently peaking between September/October across states, given the short duration of vaccine-induced immunity^39,40^. Third, our analysis provides further specificity to guide the optimal timing of the second summer-targeted vaccine dose across US states, which has limited public health guidance. Our analysis suggests that differential timing of semiannual COVID-19 waves across US states is likely driven by variation in climate, with larger winter waves in colder regions of the US leading to greater relative benefit of a fall vaccination campaign that peaks in October. In Western states, where the summer peak approaches the magnitude of the winter peak, a two-dose strategy that includes summer vaccination peaking in August and a fall dose peaking in October may provide greater benefit; if seasonal patterns further shift toward larger summer peaks, prioritizing summer-targeted vaccination could be considered.

There are several limitations to this analysis. We use a simplified model structure to describe broad patterns in SARS-CoV-2 dynamics, which does not fully account for the complexity of demography, transmission, immunity, clinical risk factors, and social contact structure that have previously been shown to affect COVID-19 disease risk^41–43^. Nevertheless, the model’s ability to capture broad differences in the timing and magnitude of outbreaks suggests that it incorporated the primary mechanistic drivers governing seasonal trends, which was the primary goal of this study. The analysis of potential vaccine impacts likely underestimates the benefit of optimal timing and schedules because it was conducted at the population level without an age- or risk-targeted model, therefore interpretation should focus on relative comparisons between states. As previously described, the inability of the model to disentangle waning immunity from viral evolution with immune evasion suggests that either or both could be the key contributor. Our model simplified the role of human behavior in influencing SARS-CoV-2 dynamics and assumed each novel variant had the same impact in the model given limited data to support more complex parameterization. Viral interactions with other pathogens, including seasonal coronaviruses and other endemic respiratory diseases, were not considered in the model structure^35^. At this stage in the pandemic, COVID-19 hospitalization data is likely the least biased surveillance data available, although it is still prone to some bias and not entirely representative of the US. Population immunity was inferred from epidemiologic data assuming relationships that may vary across individuals, variants, and immune histories. The model is broadly parameterized to represent waning immunity against infection, rather than severe disease. Finally, our study was focused on the United States, and some findings may not generalize to other settings.

## Conclusions

Our analysis found that a mechanistic model incorporating waning infection-acquired and vaccine-derived immunity, climate-dependent transmission, and variant activity effectively captured semiannual SARS-CoV-2 dynamics across US states in the post-Omicron era. These climatic relationships may help explain why colder regions experience larger winter waves, whereas states in other regions tend to show semiannual peaks shaped by humidity fluctuations. Such regional differences have broad implications for vaccine policy, particularly inclusion of a two-dose schedule and timing of vaccination uptake among high-risk groups who depend on well-timed protection.

## Methods

### Data

We calibrated our model to data from the COVID-19 Hospitalization Surveillance Network (COVID-NET), which records incidence of laboratory-confirmed COVID-19-related hospitalizations (per 100,000 persons) across US states, representing around 10% of the US population^1^. COVID-19 associated hospitalizations were defined by a positive SARS-CoV-2 test within 14 days before or during hospital admission with a clinically compatible presentation. Data was obtained for the period between January 2022 to November 2024. COVID-NET conducts surveillance data for thirteen states across the US; we model ten here due to some states having incomplete hospitalization or vaccination time series data.

To assess potential vaccine-related, climatic, genomic, and behavioral factors influencing SARS-CoV-2 transmission dynamics, we obtained data on several candidate mechanisms that may affect transmission. We used state-specific vaccination data from the US CDC^44,45^ and calculated the weekly percent of population vaccinated (see **Supplementary Material 1**). Climate data, specifically temperature and relative humidity, were obtained from Google Earth Engine^46^. We calculated the mean weekly temperature and relative humidity for each state, averaging over the specific counties that report to COVID-NET^47^. We calculated weekly shares of variant dominance^48^ using the CDC SARS-CoV-2 Variant Proportion dataset (see **Supplementary Material 1**). Lastly, we used the weekly fraction of the population that remained indoors using published county-level SafeGraph mobility data^30^ as a proxy for human behavior relevant to transmission. Additional information on how each dataset was processed can be found in **Supplementary Material 1.** This study was not human subjects research given use of publicly available secondary datasets without identifiable data.

### Modeling approach

To model patterns in SARS-CoV-2 dynamics across US states, we used a compartmental Susceptible-Exposed-Infected-Recovered-Susceptible (SEIRS) model with vaccination (V). Vaccination assumed an all-or-nothing protection mechanism. The governing equations are:

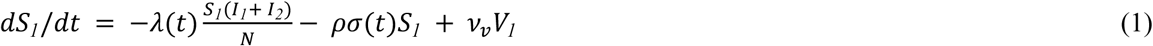

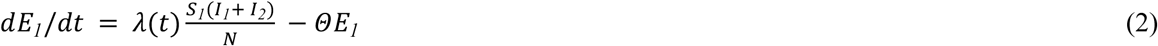

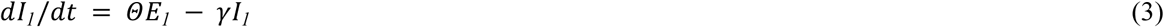

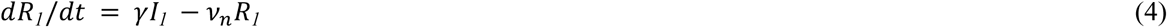

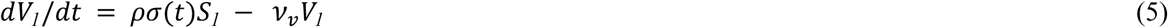

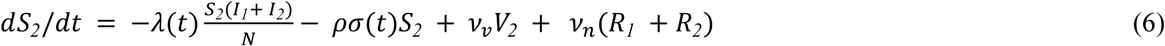

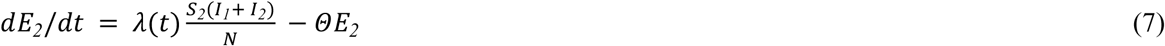

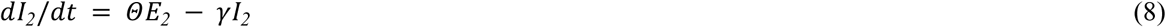

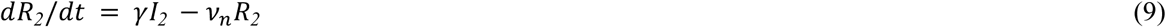

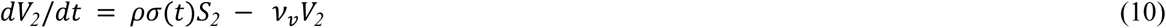

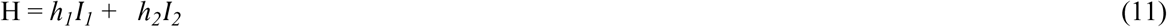

The infectious period (1/*γ*) and latent period (1/*θ*) were both set to 4 days^49–51^. The rate of vaccination *σ*(*t*) was set as the weekly percent of the population that received a single dose of a COVID-19 vaccine. Vaccine efficacy (*ρ*) against infection was set to 30% based on prior studies, and was varied in sensitivity analysis (**Supplementary Figure S2**)^39,52^. After an individual experiences their first infection, they waned into a second set of compartments, which was structured to account for the lower risk of hospitalization from subsequent SARS-CoV-2 infections due to infection-acquired protection against severe disease. The model was initialized to January 2022 conditions after the emergence of Omicron, and therefore a large fraction of the population had experienced at least one infection and were vaccinated; therefore S_1_ represents susceptibility to infection but with a majority of the population having some vaccine-derived protection against hospitalization. Alternative model structures were tested to account for vaccine-derived protection against severe disease (see **Supplementary Materials 2, Supplementary Figure S5)**. The hospitalization rates following the first (*I_1_*) and subsequent infections (*I_2_*) and the duration of immunity following infection (1/*v_n_*) and vaccination (1/*v_v_*) were calibrated, and represent a composite average across age and risk groups.

We tested seven distinct model frameworks to assess the role of infection- and vaccine-derived immunity, temperature, relative humidity, variant behavior, and human behavior patterns on transmission dynamics (**Table 1**). Our base model assumed that transmission was only influenced by waning infection- and vaccine-derived immunity (Model 1 in **Table 1**). In versions of the model that considered climatic factors (Models 2-7), the transmission rate was modulated by a negative, linear relationship with temperature (T) and/or a nonlinear, U-shaped relationship with relative humidity (RH) using representative county-level data on temperature and relative humidity. The climate relationship was globally fit across all states. We specified the functional relationship between force of infection and relative humidity and temperature as:

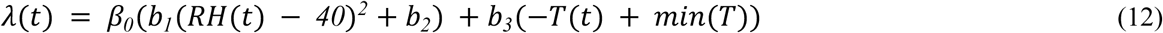

Where T(t) and RH(t) represented observed weekly climatic conditions aligning with the county locations for COVID-NET, and *b_1_*, *b_2_*, and *b_3_* were fitted parameters. In line with laboratory findings showing a linear increase in SARS-CoV-2 particle survival at cooler temperatures^14^, observed temperature T(t) was negated, and the minimum temperature observed was added to all values to ensure T(t) > 0. Laboratory studies suggest that the relationship between relative humidity and SARS-CoV-2 viral survival follows a U-shape, whereby survival is increased at both low (RH< 40%) and high (RH > 80%) relative humidity^14^. To incorporate this nonlinear relationship, we allowed for a parabolic relationship between humidity and transmission, as has been previously modeled for respiratory pathogens^3,13,53^, where impact on transmission increased both below and above *RH*(*t*) = *40%*. RH did not commonly exceed 80% across US states, and thus we did not account for high transmission probability at higher RH values in the model framework. We also fitted models that considered temperature (Model 2 in **Table 1**) and relative humidity (Model 3 in **Table 1**) separately, and performed sensitivity analysis using lagged impact (between 0-2 weeks) of climate factors on transmission.

To evaluate the impact of novel variant dynamics (Models 6-7), we defined periods of high variant circulation as periods where a novel variant circulated for a minimum of 4 weeks above 70% dominance. During these periods, we modeled immune evasion against naturally-acquired immunity with stepwise increased waning at uniform rate g(*t*), which was a calibrated term. The variants that corresponded to periods of high variant circulation were BA.5, JN.1, and XBB1.5. Several other approaches were also tested to assess the influence of variant dynamics on SARS-CoV-2 transmission, such as increased infectiousness (**Supplementary Materials 3**).

Finally, we implemented a model that considered the role of human behavioral patterns in influencing transmission in addition to other factors (Model 7 in **Table 1**). We used a time-varying parameter *B*(*t*) representing propensity for visits at indoor locations relative to outdoor locations^30^. In this version of the model, the force of infection was:

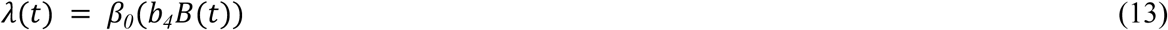

Where *b_4_* is a fitted scalar. We estimated state-specific parameters for the transmission coefficient *β_0_* and hospitalization fractions *ℎ_1_* and *ℎ_2_* given expected differences between states due to difference in age- and risk-profiles along with other factors such as healthcare access and demographics.

### Model calibration and validation

The model was calibrated in two steps using a hierarchical MCMC Metropolis Hastings algorithm with 10,000 iterations of 3 chains assuming a Poisson likelihood to compare model-estimated and observed weekly COVID-19 related hospitalizations. Parameters that would not be expected to differ across states (i.e., the durations of infection- and vaccine-derived immunity, the scalars on relative humidity, temperature, indoor activity, and variant-induced waning rate) were first fitted globally. Model convergence was assessed using *R* and posterior density plots (**Supplementary Figure 1**). A full parameter table showing fitted global parameters for the best performing model structure (Model 6) is provided in **Table 2**.

**Table 2.** Model parameters for SARS-CoV-2 mechanistic model.

| Parameter | Definition | Fit or fixed | Value (95% CrI) |
| --- | --- | --- | --- |
| $\beta_0$ | Global transmission coefficient<sup>a</sup> | Fit | 0.48 (0.44 0.52) |
| $1/\theta$ | Latent period duration (days) | Fixed<sup>52,54</sup> | 4 |
| $1/\gamma$ | Duration of infectiousness (days) | Fixed<sup>52,55</sup> | 4 |
| $1/\nu_n$ | Mean duration of infection-induced protection (months) | Fit | 4.95 (4.73, 5.19) |
| $1/\nu_v$ | Mean duration of vaccine-induced protection (months) | Fit<sup>39,40</sup> | 3.13 (2.24, 5.19) |
| $\rho$ | Vaccine effectiveness against infection<sup>b</sup> (%) | Fixed | 30 |
| $\sigma(t)$ | Vaccine coverage | Fixed | Observed |
| $h_1, h_2$ | Infection hospitalization fraction in primary and secondary infections<sup>c</sup> | Fit | 0.0095 (0.0080, 0.0111)<br>0.0030 (0.0027, 0.0032) |
| $b_1, b_2$ | Functional shape parameters on observed relative humidity | Fit | 0.0021 (0.0017, 0.0024)<br>5.97 (5.82, 6.12) |
| $b_3$ | Scalar on observed temperature | Fit | 0.058 (0.042, 0.074) |
| $g(t)$ | Multiplier on waning rate ($\nu_n$) during periods of new dominant variants | Fit | 1.25 (1.14, 1.35) |

<sup>a</sup>Model calibration included an additional state-specific transmission coefficient<sup>b</sup>Vaccine effectiveness was varied in sensitivity analysis<sup>c</sup>Infection hospitalization fraction represents a composite average of all age and risk groups
CrI; Credible interval

In a second step, we used Latin Hypercube sampling to determine state-specific parameters for the transmission term, infection hospitalization rates, and the susceptible fraction at the state level at model initialization. We use a two-step approach so that global parameters could be fitted first, and subsequent state-level differences could be accounted for while improving computational efficiency. A table describing fitted state-specific parameters is provided in **Supplementary Table S1**.

A model validation exercise was undertaken to evaluate how the final model would capture COVID-19 dynamics during the period after model calibration. We performed a model validation from December 2024 to December 2025, which was a time period not used in model calibration in the primary analysis and compared model-predicted and observed severe COVID-19 hospitalizations over time (**Supplementary Figure S3).**

### Testing relative importance of potential mechanistic drivers of SARS-CoV-2 dynamics

Informed by the best fit model, we assessed the relative importance of key epidemiological drivers affecting transmission: infection-acquired immunity (influenced by novel variants), vaccine-derived immunity, temperature, and relative humidity. To evaluate relative importance, we regressed the effective reproduction number R_e_(*t*), defined as the number of expected secondary infections in a population at time *t* given the current population immunity, against time-varying covariates, following a previously published approach^54^. R_e_(*t*) was calculated as:

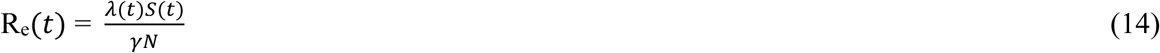

We regressed R_e_(*t*) against the observed time series for relative humidity, temperature, and the fitted fraction of the population in the recovered or vaccinated compartments (immune) for each state during the 2022-23 and 2023-24 seasons from August-February, comprising the majority of COVID-19 activity across US states. We estimated the relative contribution of these variables in rolling 5-week time periods using the Lindeman, Merenda, and Gold method^55^.

### Scenario analysis: Perturbing semiannual dynamics

We conducted scenario analyses to assess whether changes in immunity and/or viral evolution could plausibly alter future SARS-CoV-2 dynamics. We did not vary climatic conditions given that we do not expect them to deviate substantially from observed patterns in upcoming years. We used case examples from three US states (Tennessee, California, and Minnesota), chosen to represent diverse SARS-CoV-2 dynamics across the US, using the 2023-24 season. We varied the mean duration of immunity (20-52 weeks), which is meant to also represent a changing rate in immune-evading viral evolution. In these simulations, the perturbed parameters were set to new values in January 2023, and projections were simulated until June 2024.

### Scenario analysis: Optimizing vaccination timing and schedule

We evaluated the impact of vaccine uptake timing and different vaccine dosing strategies on COVID-19 hospitalization burden across US states using the 2023-2024 season as a case example. First, we shifted the observed vaccination curve between 6 weeks earlier and 6 weeks later to assess whether there may be more optimal vaccination timing across states. Then, we tested three vaccine dose schedules (annual fall dose, annual summer dose, two-dose in fall and summer) and estimated projected hospitalizations between the period of June 2023-June 2024. We simulated hypothetical vaccine uptake curves assuming cumulative vaccine coverage based on observed state-specific coverage levels in the winter season (ranging from 9-27% coverage across states); the two-dose strategy assumed equal coverage for both doses. We compared the predicted hospitalizations to a counterfactual with no vaccination. We also assessed scenarios with more optimistic vaccination coverage of 50% across states (**Supplementary Figure S4**); more detail on this approach is in **Supplementary Materials 4**.

## Data Availability

All data in the present work are available at: https://github.com/sbents/covid_seasonality

## Code sharing statement

All code used for this analysis and data has been made publicly available at: https://github.com/sbents/covid_seasonality

## Acknowledgments

We acknowledge those involved in data collection of these public datasets. The study content is solely the responsibility of the authors and does not necessarily represent the official views of the National Institutes of Health.

## Author contributions

Ms. Samantha Bents and Dr. Nathan Lo had full access to all the data in the study and take responsibility for the integrity of the data and the accuracy of the data analysis.

Study concept and design: SJB, NCL

Statistical analysis: SJB, NCL

Analytic coding: SJB

Acquisition, analysis, or interpretation of data: All authors

First draft of the manuscript: SJB, NCL

Critical revision of the manuscript: All authors

Contributed intellectual material and approved final draft: All authors

## References

1. Coronavirus Disease 2019 (COVID-19) Hospitalization Surveillance Network (COVID-NET). CDC.

2. Baker, R. E. et al. Epidemic dynamics of respiratory syncytial virus in current and future climates. Nat. Commun. 10, 5512 (2019).

3. Shaman, J., Pitzer, V. E., Viboud, C., Grenfell, B. T. & Lipsitch, M. Absolute Humidity and the Seasonal Onset of Influenza in the Continental United States. PLoS Biol. 8, e1000316 (2010).

4. Viboud, C. et al. Synchrony, Waves, and Spatial Hierarchies in the Spread of Influenza. Science 312, 447–451 (2006).

5. Dalziel, B. D. et al. Urbanization and humidity shape the intensity of influenza epidemics in U.S. cities. Science 362, 75–79 (2018).

6. Kimball, P. et al. Urban contact patterns shape respiratory syncytial virus epidemics with implications for vaccination. Sci. Adv. 11, eady5457 (2025).

7. Carabelli, A. M. et al. SARS-CoV-2 variant biology: immune escape, transmission and fitness. Nat. Rev. Microbiol. https://doi.org/10.1038/s41579-022-00841-7 (2023) doi:10.1038/s41579-022-00841-7.

8. Mykytyn, A. Z., Fouchier, R. A. & Haagmans, B. L. Antigenic evolution of SARS coronavirus 2. Curr. Opin. Virol. 62, 101349 (2023).

9. Koelle, K., Martin, M. A., Antia, R., Lopman, B. & Dean, N. E. The changing epidemiology of SARS-CoV-2. Science 375, 1116–1121 (2022).

10. Chemaitelly, H. et al. Duration of immune protection of SARS-CoV-2 natural infection against reinfection. J. Travel Med. 29, taac109 (2022).

11. Stein, C. et al. Past SARS-CoV-2 infection protection against re-infection: a systematic review and meta-analysis. The Lancet 401, 833–842 (2023).

12. Shaman, J. & Kohn, M. Absolute humidity modulates influenza survival, transmission, and seasonality. Proc. Natl. Acad. Sci. 106, 3243–3248 (2009).

13. Mahmud, A. S., Martinez, P. P. & Baker, R. E. The impact of current and future climates on spatiotemporal dynamics of influenza in a tropical setting. PNAS Nexus 2, pgad307 (2023).

14. Morris, D. H. et al. Mechanistic theory predicts the effects of temperature and humidity on inactivation of SARS-CoV-2 and other enveloped viruses. eLife 10, e65902 (2021).

15. Baker, R. E., Yang, W., Vecchi, G. A., Metcalf, C. J. E. & Grenfell, B. T. Susceptible supply limits the role of climate in the early SARS-CoV-2 pandemic. Science 369, 315–319 (2020).

16. Carlson, C. J., Gomez, A. C. R., Bansal, S. & Ryan, S. J. Misconceptions about weather and seasonality must not misguide COVID-19 response. Nat. Commun. 11, 4312 (2020).

17. Chinazzi, M. et al. The effect of travel restrictions on the spread of the 2019 novel coronavirus (COVID-19) outbreak. Science 368, 395–400 (2020).

18. Perofsky, A. C. et al. Impacts of human mobility on the citywide transmission dynamics of 18 respiratory viruses in pre- and post-COVID-19 pandemic years. Nat. Commun. 15, 4164 (2024).

19. Ryu, S., Ali, S. T., Cowling, B. J. & Lau, E. H. Y. Effects of School Holidays on Seasonal Influenza in South Korea, 2014–2016. J. Infect. Dis. 222, 832–835 (2020).

20. Martin, L. J. et al. Influenza-like illness-related emergency department visits: Christmas and New Year holiday peaks and relationships with laboratory-confirmed respiratory virus detections, Edmonton, Alberta, 2004–2014. Influenza Other Respir. Viruses 11, 33–40 (2017).

21. Ewing, A., Lee, E. C., Viboud, C. & Bansal, S. Contact, travel, and transmission: The impact of winter holidays on influenza dynamics in the United States. J. Infect. Dis. jiw642 (2016) doi:10.1093/infdis/jiw642.

22. Roper, L. E. et al. Use of Additional Doses of 2024–2025 COVID-19 Vaccine for Adults Aged ≥65 Years and Persons Aged ≥6 Months with Moderate or Severe Immunocompromise: Recommendations of the Advisory Committee on Immunization Practices — United States, 2024. MMWR Morb. Mortal. Wkly. Rep. 73, 1118–1123 (2024).

23. Du, Y., Paritala, S., Xu, Y., Maloney, P. & Lin, D.-Y. Durability of 2024-2025 COVID-19 Vaccines Against JN.1 Subvariants. JAMA Intern. Med. 185, 1501 (2025).

24. Bobrovitz, N. et al. Protective effectiveness of previous SARS-CoV-2 infection and hybrid immunity against the omicron variant and severe disease: a systematic review and meta-regression. Lancet Infect. Dis. 23, 556–567 (2023).

25. Ssentongo, P. et al. SARS-CoV-2 vaccine effectiveness against infection, symptomatic and severe COVID-19: a systematic review and meta-analysis. BMC Infect. Dis. 22, 439 (2022).

26. Peng, S. et al. Stability of SARS-CoV-2 in cold-chain transportation environments and the efficacy of disinfection measures. Front. Cell. Infect. Microbiol. 13, 1170505 (2023).

27. Casanova, L. M., Jeon, S., Rutala, W. A., Weber, D. J. & Sobsey, M. D. Effects of Air Temperature and Relative Humidity on Coronavirus Survival on Surfaces. Appl. Environ. Microbiol. 76, 2712–2717 (2010).

28. Kudo, E. et al. Low ambient humidity impairs barrier function and innate resistance against influenza infection. Proc. Natl. Acad. Sci. 116, 10905–10910 (2019).

29. Prather, K. A., Wang, C. C. & Schooley, R. T. Reducing transmission of SARS-CoV-2. Science 368, 1422–1424 (2020).

30. Susswein, Z., Rest, E. C. & Bansal, S. Disentangling the rhythms of human activity in the built environment for airborne transmission risk: An analysis of large-scale mobility data. eLife 12, e80466 (2023).

31. Rowe, B. R., Canosa, A., Drouffe, J. M. & Mitchell, J. B. A. Simple quantitative assessment of the outdoor versus indoor airborne transmission of viruses and COVID-19. Environ. Res. 198, 111189 (2021).

32. Shah, M. M. et al. Seasonality of Common Human Coronaviruses, United States, 2014–20211. Emerg. Infect. Dis. 28, 1970–1976 (2022).

33. Lee, S. S., Viboud, C. & Petersen, E. Understanding the rebound of influenza in the post COVID-19 pandemic period holds important clues for epidemiology and control. Int. J. Infect. Dis. 122, 1002–1004 (2022).

34. Anastasiou, O. E. et al. Seasonality of Non-SARS, Non-MERS Coronaviruses and the Impact of Meteorological Factors. Pathogens 10, 187 (2021).

35. Kissler, S. M., Tedijanto, C., Goldstein, E., Grad, Y. H. & Lipsitch, M. Projecting the transmission dynamics of SARS-CoV-2 through the postpandemic period. Science 368, 860–868 (2020).

36. Wells, C. R. et al. Evaluation of Strategies for Transitioning to Annual SARS-CoV-2 Vaccination Campaigns in the United States. Ann. Intern. Med. 177, 609–617 (2024).

37. Park, H. J. et al. Comparing frequency of booster vaccination to prevent severe COVID-19 by risk group in the United States. Nat. Commun. 15, 1883 (2024).

38. Healthcare Professionals: Adult Immunization Schedule by Age.

39. Lau, J. J. et al. Real-world COVID-19 vaccine effectiveness against the Omicron BA.2 variant in a SARS-CoV-2 infection-naive population. Nat. Med. 29, 348–357 (2023).

40. Link-Gelles, R. et al. Early Estimates of Updated 2023–2024 (Monovalent XBB.1.5) COVID-19 Vaccine Effectiveness Against Symptomatic SARS-CoV-2 Infection Attributable to Co-Circulating Omicron Variants Among Immunocompetent Adults — Increasing Community Access to Testing Program, United States, September 2023–January 2024. MMWR Morb. Mortal. Wkly. Rep. 73, 77–83 (2024).

41. Aburto, J. M., Tilstra, A. M., Floridi, G. & Dowd, J. B. Significant impacts of the COVID-19 pandemic on race/ethnic differences in US mortality. Proc. Natl. Acad. Sci. 119, e2205813119 (2022).

42. Acosta, N. et al. Longitudinal SARS-CoV-2 RNA wastewater monitoring across a range of scales correlates with total and regional COVID-19 burden in a well-defined urban population. Water Res. 220, 118611 (2022).

43. Tisminetzky, M. et al. Age, Multiple Chronic Conditions, and COVID-19: A Literature Review. J. Gerontol. Ser. A 77, 872–878 (2022).

44. Weekly COVID-19 Vaccination Dashboard. COVIDVaxView.

45. Monthly Cumulative Number and Percent of Persons Who Received 1+ COVID-19 Vaccination Doses, by Season, Age Group, and Jurisdiction, United States. National Center for Immunization and Respiratory Diseases (NCIRD).

46. Gorelick, N., et al. Google Earth Engine: Planetary-scale geospatial analysis for everyone. Remote Sens. Environ. 202, 18–27 (2017).

47. Garg, S. et al. Hospitalization Rates and Characteristics of Patients Hospitalized with Laboratory-Confirmed Coronavirus Disease 2019 — COVID-NET, 14 States, March 1–30, 2020. MMWR Morb. Mortal. Wkly. Rep. 69, 458–464 (2020).

48. SARS-CoV-2 Variant Proportions. CORVD Laboratory Branch.

49. Chen, D. et al. Inferring time-varying generation time, serial interval, and incubation period distributions for COVID-19. Nat. Commun. 13, 7727 (2022).

50. Wu, Y. et al. Incubation Period of COVID-19 Caused by Unique SARS-CoV-2 Strains: A Systematic Review and Meta-analysis. JAMA Netw. Open 5, e2228008 (2022).

51. Puhach, O., Meyer, B. & Eckerle, I. SARS-CoV-2 viral load and shedding kinetics. Nat. Rev. Microbiol. https://doi.org/10.1038/s41579-022-00822-w (2022) doi:10.1038/s41579-022-00822-w.

52. Feldstein, L. R. et al. Effectiveness of mRNA COVID-19 Vaccines and Hybrid Immunity in Preventing SARS-CoV-2 Infection and Symptomatic COVID-19 Among Adults in the United States. J. Infect. Dis. 231, e743–e753 (2025).

53. Shaman, J. & Kohn, M. Absolute humidity modulates influenza survival, transmission, and seasonality. Proc. Natl. Acad. Sci. 106, 3243–3248 (2009).

54. Te Beest, D. E., Van Boven, M., Hooiveld, M., Van Den Dool, C. & Wallinga, J. Driving Factors of Influenza Transmission in the Netherlands. Am. J. Epidemiol. 178, 1469–1477 (2013).

55. Grömping, U. Relative Importance for Linear Regression in R: The Package relaimpo. J. Stat. Softw. 17, (2006).

