## Supplementary for "Interplay of Immunity, Climate, and Viral Evolution Explains Semiannual SARS-CoV-2 Dynamics with Implications for Control"

### Material and Methods

#### 1. Additional information on methods for data processing

COVID-NET reports a weekly COVID-19 associated hospitalization incidence (per 100,00 persons), which we transformed to absolute weekly counts for each state by scaling it to the state's population based on the 2023 census[1]. A COVID-19-associated hospitalization is defined by a positive SARS-CoV-2 test within 14 days before or during hospital admission with a clinically compatible presentation.

For vaccination data up until 2024, we used the CDC CovidVax dataset[2] for state-level vaccination data. Data was processed to calculate weekly vaccination rates. Incident vaccinations were computed as the difference in cumulative doses administered between consecutive time points, then normalized by state population size (2023 Census estimates) to obtain weekly vaccination rates.

For the 2024-25 vaccination data, we sourced monthly observed vaccination uptake data from the CDC[3]. The data provides the cumulative number of doses administered across US states at the monthly timescale. From this, we calculated monthly incident doses administered and then disaggregated this to a weekly timescale. To transform this into the proportion of the population that received vaccination on the weekly timescale, we divided weekly incident doses administered by the population size of the state, ensuring overall vaccination coverage levels were preserved.

Climate data for relative humidity and temperature were sourced from Google Earth Engine (GEE)[4]. We calculated the mean weekly temperature and relative humidity for each state, averaging over the specific counties that report to the COVID-NET surveillance program[1]. This ensured that the climate exposures implemented in the model were derived from the geographical area that reported COVID-19-associated hospitalizations, which is particularly important for larger US states where there is substantial variation in climate across the state. Using GEE, we extracted ERA5-Land hourly temperature and dewpoint data and aggregated it into weekly averages. From temperature and dewpoint data, the vapor pressure deficit is calculated and then converted to relative humidity (%) using a standard meteorological formula (Magnus approximation).

We considered the role of variant activity by sourcing CDC SARS-CoV-2 Variant Proportion dataset, and calculated shares of variant dominance. This dataset provided the biweekly proportion of each circulating variant that was observed from hospital-based samples across the Department of Health and Human Service regions[5]: New York (Region 2- New York City), Georgia, Tennessee (Region 4- Atlanta), Michigan, Minnesota, Ohio, (Region 5- Chicago), Utah, Colorado (Region 8- Denver), California (Region 9- San Francisco), Oregon (Region 10- Seattle). Individual lineages were grouped into broader variant classifications: BA.1.1 (BA.1.1, B.1.1.529), BQ (BQ.1, BQ.1.1), BA.2 (BA.2, BA.2.12.1), BA.4/5 (BA.4, BA.5), XBB (XBB.1.5, XBB.1.9.1, XBB.1.16, XBB.2.3), EG/HV (EG.5, HV.1), JN.1 (JN.1, JN.1.7), KP/LB (KP.2, KP.2.3, KP.3, KP.3.1.1, LB.1), and a residual low-circulating variants category (comprised < 10% in a given region-time period). To obtain weekly shares of variant dominance, we disaggregated the biweekly time series by interpolating a week's share as the average and current lagged biweekly values.

Human behavior was tested as a candidate model covariate using previously processed state-level SafeGraph data[6]. Briefly, SafeGraph Points of Interest (POIs) across the United States, were categorized using six-digit NAICS codes as primarily indoor, outdoor, or unclear. Using weekly county-level visit counts, the indoor activity metric was defined as the ratio of normalized indoor to outdoor visits, which was then mean-centered at the county level. We subsetted this dataset to only include counties that report to COVID-NET and averaged across these counties to obtain representative means at the state level.

#### 2. Alternative model structure

We implemented an alternative model structure where individuals waned from $V_1$ to $S_2$ instead of $S_1$, representing stronger protection from the first vaccination event against hospitalization. Model equations are provided below:

$$dS_1/dt = -\lambda(t)\frac{S_1(I_1+I_2)}{N} - \rho\sigma(t)S_1 \quad (1)$$

$$dE_1/dt = \lambda(t)\frac{S_1(I_1+I_2)}{N} - \Theta E_1 \quad (2)$$

$$dI_1/dt = \Theta E_1 - \gamma I_1 \quad (3)$$

$$dR_1/dt = \gamma I_1 - \nu_n R_1 \quad (4)$$

$$dV_1/dt = \rho\sigma(t)S_1 - \nu_v V_1 \quad (5)$$

$$dS_2/dt = -\lambda(t)\frac{S_2(I_1+I_2)}{N} - \rho\sigma(t)S_2 + \nu_v V_2 + \nu_n(R_1 + R_2) + \nu_v V_1 \quad (6)$$

$$dE_2/dt = \lambda(t)\frac{S_2(I_1+I_2)}{N} - \Theta E_2 \quad (7)$$

$$dI_2/dt = \Theta E_2 - \gamma I_2 \quad (8)$$

$$dR_2/dt = \gamma I_2 - \nu_n R_2 \quad (9)$$

$$dV_2/dt = \rho\sigma(t)S_2 - \nu_v V_2 \quad (10)$$

$$H = h_1 I_1 + h_2 I_2 \quad (11)$$

In this alternative model structure, we found that the model fit a larger portion of the population in the initial S1 compartment, and otherwise estimated similar epidemiological parameters. We assessed the number of predicted hospitalizations under vaccination scenarios using this alternative model structure and found similar projected impacts, with regional patterns in vaccination impact conserved (**Figure S5**).

#### 3. Methods for incorporating variants

We tested three separate approaches to incorporate variant-level data into the modeling framework for SARS-CoV-2 transmission dynamics based on biologically plausible effects. Variants were represented using the weekly proportion of total samples attributed to each circulating variant (See Supplementary 1 for additional information on data processing).

Following the assumption that increased transmissibility of SARS-CoV-2 should be observed during periods when individual variants dominated the variant landscape[7], we calculated a metric that represented relative variant frequency dominance. In every week, the relative frequency dominance was calculated as:

$$f(t) \ = max(p_v(t))/(1 \ - \ max(p_v(t)))$$

where $p_v(t)$ represents the proportion $p$ of a given variant *v* at time *t*.

We then modeled variants with the following approaches:

1. *Stepwise waning in infection-acquired immunity associated with high variant activity*

   We allowed variants to influence the mean duration of time for which individuals are in the Recovered compartments following infection, mimicking the process of immune evasion, when an antigenically novel variant renders pre-existing infection-acquired immunity less protective against a new infection. In this model specification, we allowed for a stepwise increase in the rate of waning following natural infection associated with periods of high variant activity.

   We used a stringent definition for periods of high variant activity, where the estimated rate of waning would increase (via a fitted multiplier) when the frequency dominance of a variant was increasing ($f(t) > 0$), but only for major variants that circulated for a minimum of 1 month above 70% dominance, corresponding to the BA.5, JN.1, and XBB1.5 variants (see Methods). This was the version used in the primary model in the study.

2. *Waning infection-acquired immunity with variant activity*

We also tested a less stringent approach, where we defined high variant activity as weeks for which the slope of frequency dominance ($f(t)$) was positive, only for variants that were able to reach 70% of total samples throughout the study period, regardless of the duration of weeks they circulated for. Five variants met this definition across states. We allowed for a stepwise increase in the rate of waning, fitted as a time-varying multiplier on the fitted duration of immunity, during the weeks where $f(t)$ was positive. This approach led to reduced model performance compared to the first approach, as measured by log-likelihood.

3. *Increasing transmissibility in line with time-varying variant dominance*

We also tested an approach where variant activity could more gradually influence the transmission term (representing increased infectiousness associated with novel variants), instead of the waning rate. We then allowed for $f(t)$ to influence the transmission term at all time points in the modeling framework so that:

$$\lambda(t) = \beta_0(b_1(RH(t) - 40)^2 + b_2) + b_3(-T(t) + min(T) + b_5(f(t)\ )$$

where $b_5$ represents a scalar on time-varying relative frequency dominance. Relative to the first approach, this model formulation also performed suboptimally (as measured by log-likelihood), and fitted $b_5$ to values near 0.

#### 4. Assessing impact of fall, summer, and two-dose vaccine schedule

We simulated three potential vaccination strategies and assessed impact across states in counterfactual scenarios for the 2023-24 season. Using state-level observed annual vaccination uptake levels in the 2023-2024 season[3], we simulated incident vaccination curves V($t$):

$$V(t) = \frac{K}{1 + e^{r(t - t_0)}}$$

Where K = state-specific observed cumulative vaccination coverage, $t_0$ is the peak of vaccination, and the growth rate $r$ is fixed at .6. We set the peak of vaccination under the summer and fall strategy to be one month before the average semiannual peak across states, corresponding to weeks 32 and 43. In the two-dose vaccination strategy, vaccination peaked at both weeks 32 and 43, and total cumulative coverage was reached for both doses. We additionally assessed more optimistic vaccination coverage levels, where K = .50 for all states, corresponding to 50% of the population vaccinated. Results shown below, which demonstrate greater overall vaccination impact but similar regional trends in the impact of the summer, fall, and two-dose vaccination strategy.

### Figures and Tables

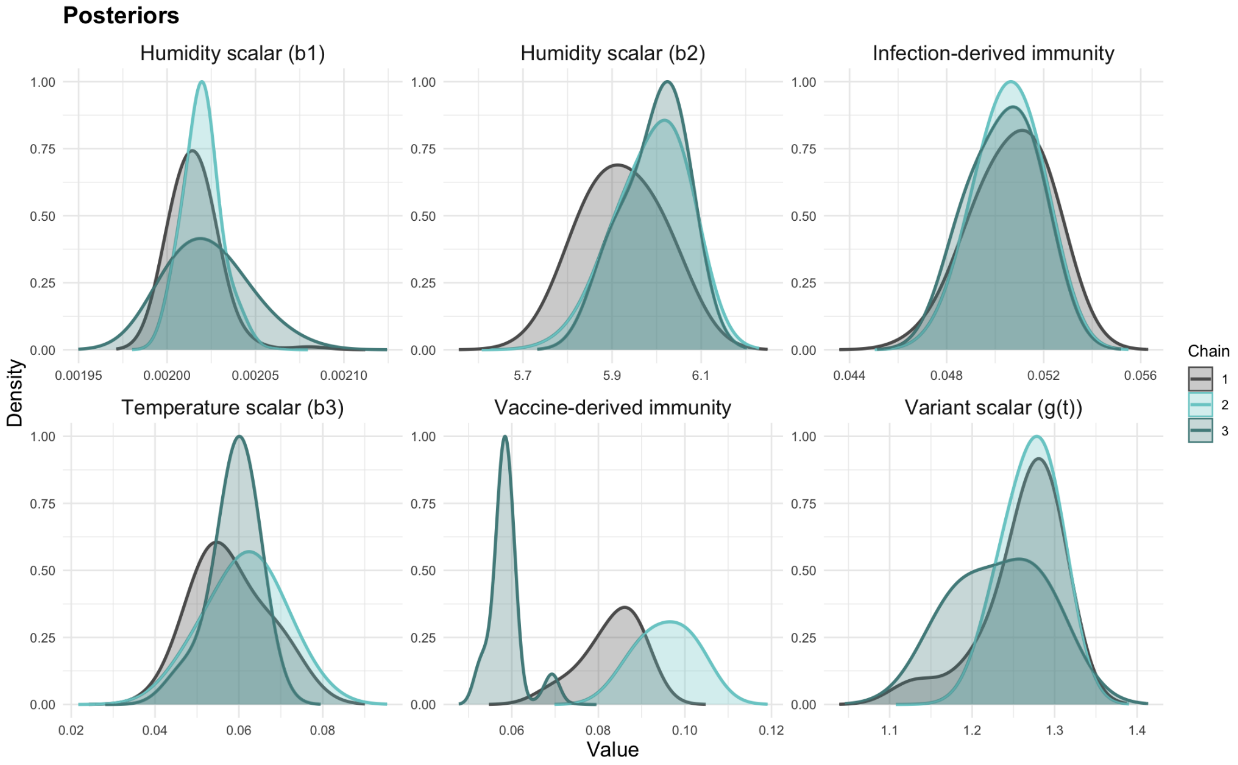


**Figure S1. Posterior distribution of globally fitted model parameters.** Density plots are shown for the posteriors of each parameter fitted in the global MCMC calibration process, including the transmission coefficient, scalars on humidity and temperature, the variant scalar, and the duration of infection- and vaccine-derived immunity. Density plots are colored by chain for the post-burn-in period, showing general posterior agreement.

**Table S1. State-specific parameters.** State-specific parameters for the transmission coefficient ($\beta_0$), initial susceptible fraction (S0), and hospitalization fractions following the first and second infections ($h_1$ and $h_2$). These parameters were fitted using Latin Hypercube sampling independently for each state; states shared global parameters including scalars on climate factors, duration of protection from infection and vaccination, and time-varying variant multiplier.

| Location | $\beta_0$ | $h_1$ | $h_2$ | S0 |
|---|---|---|---|---|
| Waning immunity, temperature, relative humidity | | | | |
| CA | 0.62 | 0.0067 | 0.0026 | 0.37 |
| CO | 0.43 | 0.0051 | 0.0023 | 0.69 |
| GA | 0.45 | 0.0065 | 0.0028 | 0.36 |
| MI | 0.44 | 0.0115 | 0.0023 | 0.36 |
| MN | 0.38 | 0.0128 | 0.0023 | 0.52 |
| NY | 0.36 | 0.0158 | 0.0037 | 0.56 |
| OH | 0.38 | 0.0080 | 0.0024 | 0.55 |
| OR | 0.52 | 0.0107 | 0.0016 | 0.25 |
| TN | 0.47 | 0.0073 | 0.0017 | 0.38 |
| UT | 0.56 | 0.0087 | 0.0011 | 0.24 |

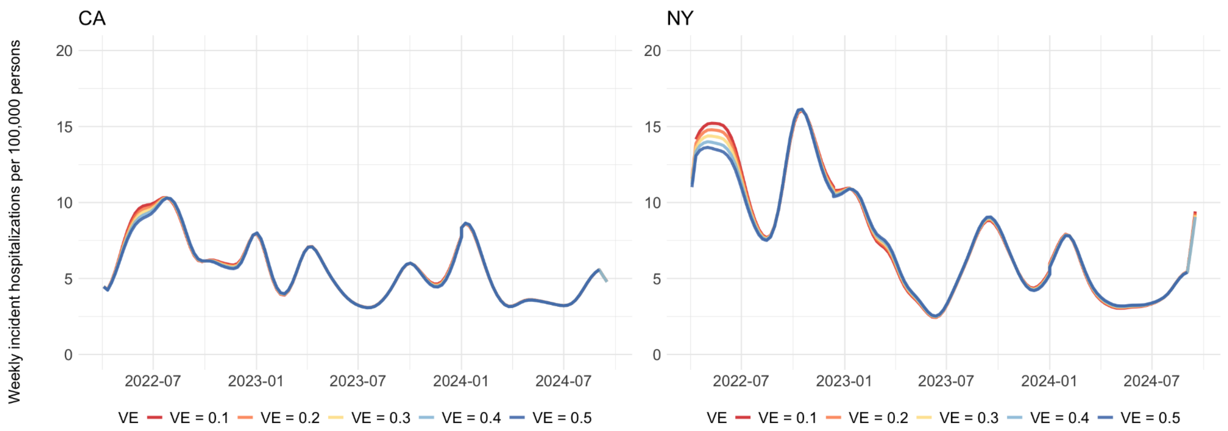


**Figure S2. One-way sensitivity analyses for vaccine effectiveness.** Vaccine effectiveness was varied between 10 and 50%, showing a limited influence on disease dynamics across states (California and New York shown).

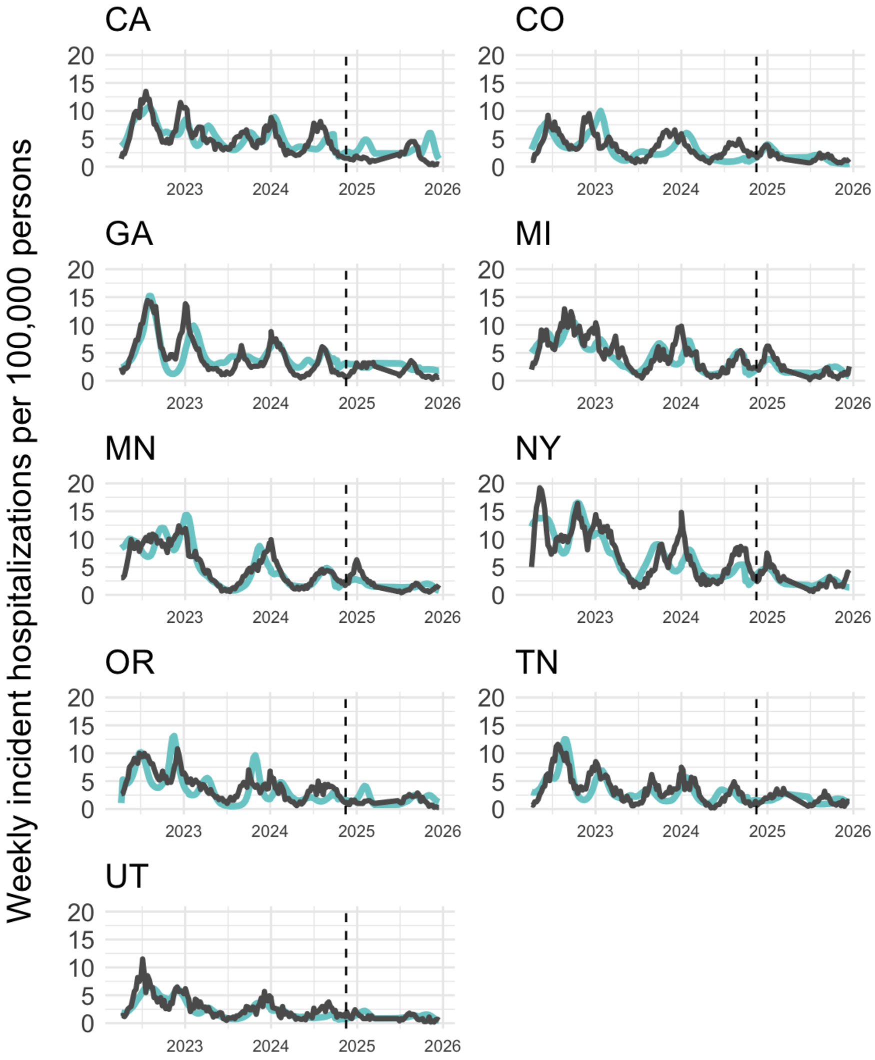


**Figure S3. Model validation over the period of December 2024 to December 2026.** Using the best performing model, we projected simulations forward to assess model predictions for December 2024-December 2025 against observed hospitalizations, incorporating historical temperature and relative humidity. We assumed that the risk of hospitalization was 60% that of the prior season, in line with estimates of continued attenuation of severe disease[8]. Variants were not considered in the validation period, given that none circulated above 70% dominance for a minimum of 4 weeks (definition used in original model fit). We found that the model generally projected observed peaks during the validation period across states.

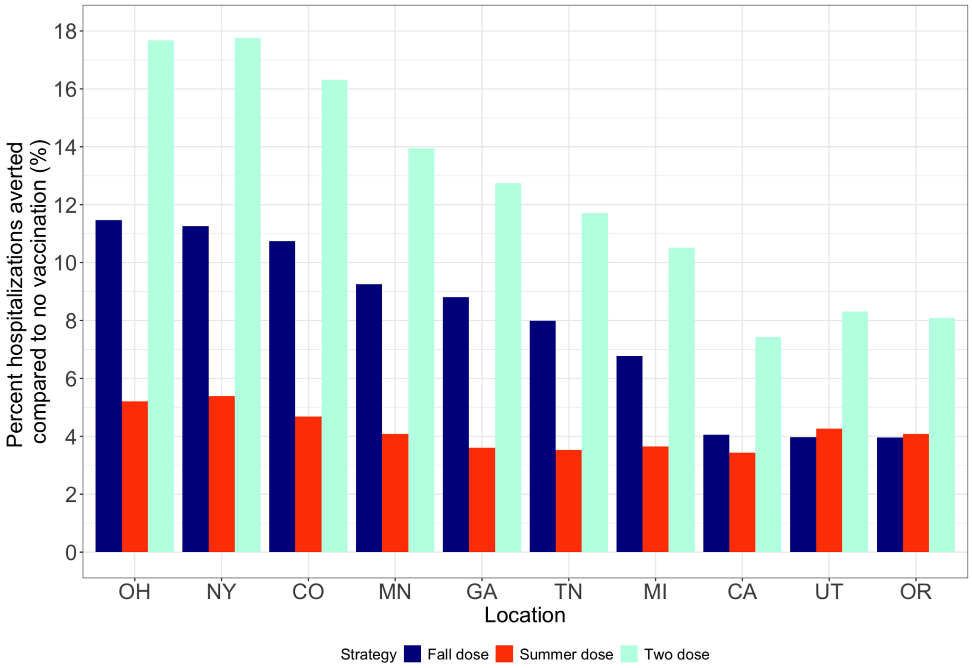


**Figure S4. Predicted impact of different vaccine schedules assuming optimistic coverage.** Predicted change in hospitalizations for a summer, fall, and combined summer and fall vaccination approach, relative to a no vaccination scenario using an optimistic vaccine coverage (sensitivity analysis for Figure 4D). Here, we assumed optimistic annual vaccine coverage levels of 50% for every state, finding that while regional trends were conserved, vaccine impact was larger across states.

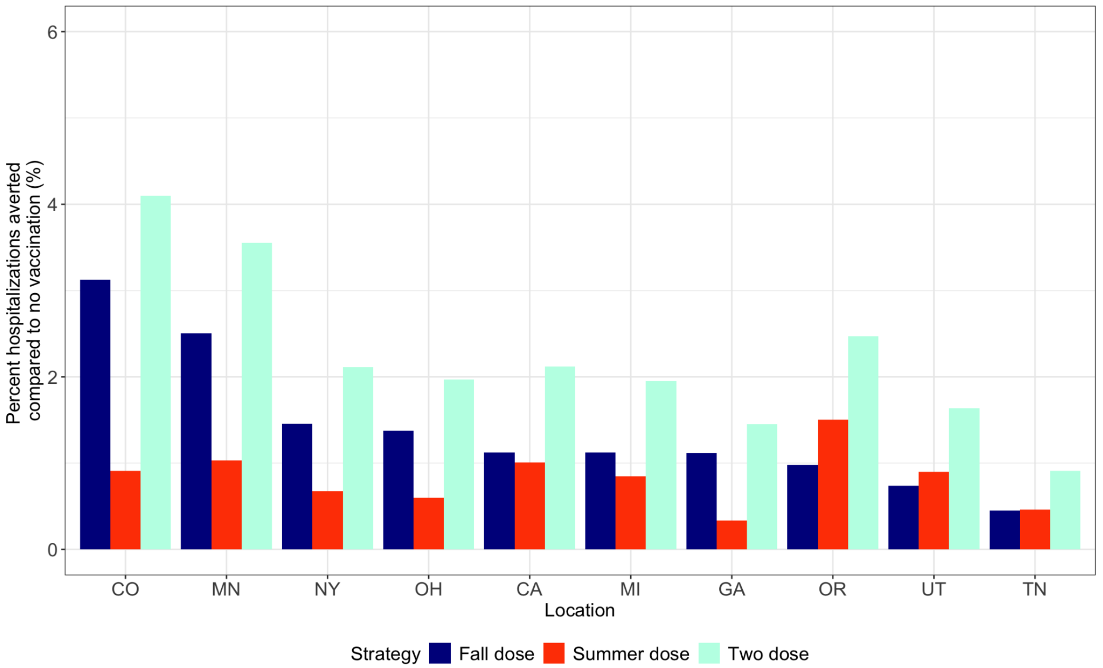


**Figure S5. Predicted vaccination impact using alternative model structure.** We conducted a model structure sensitivity analysis (described in Supplementary Materials 2) in which the first vaccination event is associated with a lower risk of hospitalization by waning into the S2 compartment with a lower rate of hospitalization upon future infection. Using this alternative model structure, we found the predicted impact of summer, fall, or two-dose vaccination strategy compared to a no-vaccination strategy was similar to the model structure presented in the main analysis (Figure 4D).

#### Supplementary References


1. Coronavirus Disease 2019 (COVID-19) Hospitalization Surveillance Network (COVID-NET). CDC.
2. Weekly COVID-19 Vaccination Dashboard. COVIDVaxView.
3. Monthly Cumulative Number and Percent of Persons Who Received 1+ COVID-19 Vaccination Doses, by Season, Age Group, and Jurisdiction, United States. National Center for Immunization and Respiratory Diseases (NCIRD).
4. Gorelick, N. *et al.* Google Earth Engine: Planetary-scale geospatial analysis for everyone. *Remote Sens. Environ.* **202**, 18–27 (2017).
5. SARS-CoV-2 Variant Proportions. CORVD Laboratory Branch.
6. Susswein, Z., Rest, E. C. & Bansal, S. Disentangling the rhythms of human activity in the built environment for airborne transmission risk: An analysis of large-scale mobility data. *eLife* **12**, e80466 (2023).
7. Figgins, M. D. & Bedford, T. Frequency dynamics predict viral fitness, antigenic relationships and epidemic growth. Preprint at https://doi.org/10.1101/2024.12.02.24318334 (2024).
8. Flisiak, R. *et al.* Differences in the Clinical Course of COVID-19 in Patients Hospitalized in the 2023/2024 and 2024/2025 Seasons. *J. Clin. Med.* **14**, 5992 (2025).
